# Mitochondrial DNA variant detection in over 6,500 rare disease families by the systematic analysis of exome and genome sequencing data resolves undiagnosed cases

**DOI:** 10.1101/2024.12.22.24319370

**Authors:** Sarah L. Stenton, Kristen Laricchia, Nicole J. Lake, Sushma Chaluvadi, Vijay Ganesh, Stephanie DiTroia, Ikeoluwa Osei-Owusu, Lynn Pais, Emily O’Heir, Christina Austin-Tse, Melanie O’Leary, Mayada Abu Shanap, Chelsea Barrows, Seth Berger, Carsten G. Bönnemann, Kinga M. Bujakowska, Dean R. Campagna, Alison G. Compton, Sandra Donkervoort, Mark D. Fleming, Lyndon Gallacher, Joseph G. Gleeson, Goknur Haliloglu, Eric A. Pierce, Emily M. Place, Vijay G. Sankaran, Akiko Shimamura, Zornitza Stark, Tiong Yang Tan, David R. Thorburn, Susan M. White, Genomics Research to Elucidate the Genetics of Rare Diseases (GREGoR) Consortium, Eric Vilain, Monkol Lek, Heidi L. Rehm, Anne O’Donnell-Luria

**Affiliations:** Program in Medical and Population Genetics, Broad Institute of MIT and Harvard, Cambridge, MA USA; Division of Genetics and Genomics, Boston Children’s Hospital, Harvard Medical School, Boston, MA, USA; Department of Genetics, Yale School of Medicine, New Haven, CT, USA; Center for Genomic Medicine, Massachusetts General Hospital, Boston, MA, USA; Hematology/Oncology, Bone Marrow Transplantation and Cellular Therapy, Pediatric Department, King Hussein Cancer Centre (KHCC), Amman, Jordan; University of California, Department of Neurosciences, San Diego, CA; Rady Children’s Institute for Genomic Medicine, San Diego, CA, USA; Children’s National Research Institute, Washington, DC, USA; Neuromuscular and Neurogenetic Disorders of Childhood Section, National Institute of Neurological Disorders and Stroke, National Institutes of Health, Bethesda, MD, USA; Ocular Genomics Institute, Massachusetts Eye and Ear, Department of Ophthalmology, Harvard Medical School, Boston, MA, USA; Department of Pathology, Boston Children’s Hospital and Harvard Medical School, Boston, Massachusetts, USA; Victorian Clinical Genetics Services, Murdoch Children’s Research Institute, Flemington Road, Melbourne, Australia; Department of Paediatrics, University of Melbourne, Melbourne, Australia; Division of Hematology/Oncology, Boston Children’s Hospital, Harvard Medical School, Boston, MA 02115, USA; Department of Pediatric Oncology, Dana-Farber Cancer Institute, Harvard Medical School, Boston, MA 02215, USA; Howard Hughes Medical Institute, Boston, MA 02115, USA; Broad Institute of MIT and Harvard, Cambridge, MA 02142, USA; Department of Hematology and Oncology, Boston Children’s Hospital, Boston, MA, USA; Institute for Clinical and Translational Science, University of California, Irvine, CA, USA

## Abstract

**Background:** Variants in the mitochondrial genome (mtDNA) cause a diverse collection of mitochondrial diseases and have extensive phenotypic overlap with Mendelian diseases encoded on the nuclear genome. The mtDNA is often not specifically evaluated in patients with suspected Mendelian disease, resulting in overlooked diagnostic variants.

**Methods:** Using dedicated pipelines to address the technical challenges posed by the mtDNA - circular genome, variant heteroplasmy, and nuclear misalignment - single nucleotide variants, small indels, and large mtDNA deletions were called from exome and genome sequencing data, in addition to RNA-sequencing when available. A cohort of 6,660 rare disease families were analyzed (5,625 genetically undiagnosed, 84%) from the Genomics Research to Elucidate the Genetics of Rare diseases (GREGoR) Consortium as well as other rare disease cohorts.

**Results:** Diagnostic mtDNA variants were identified in 10 previously genetically undiagnosed families (one large deletion, eight reported pathogenic variants, one novel pathogenic variant). In one additional undiagnosed proband, the detection of >900 heteroplasmic variants provided functional evidence of pathogenicity to a novel *de novo* variant in the nuclear gene *POLG* (DNA polymerase gamma), responsible for mtDNA replication and repair.

**Conclusion:** mtDNA variant calling from data generated by exome and genome sequencing for nuclear variant analysis resulted in a genetic diagnosis or detection of a candidate variant for 0.4% of undiagnosed families affected by a broad range of rare diseases.

## INTRODUCTION

Mitochondrial diseases (MDs) result from impaired cellular energy metabolism due to defects in the mitochondrial organelle.^1^ Amongst rare genetic diseases, MDs are prime examples of the diagnostic challenge faced by geneticists, given vast genetic heterogeneity, with dual encoding on the nuclear and mitochondrial genome (mtDNA), and broad spectrum of associated clinical manifestations.^2^

The mtDNA is a circular, 16,569 base pair, double-stranded DNA molecule present in hundreds to thousands of copies per cell. It encodes 13 protein-coding genes, 22 transfer RNA (tRNA) genes, and two ribosomal RNA (rRNA) genes, that are essential to mtDNA function. Pathogenic variants in the mtDNA are responsible for approximately 75% of adult-onset and 20-25% of pediatric-onset MD.^3^ They span single nucleotide variants (SNVs), small insertions/deletions (indels), and large mtDNA deletions, and are estimated to cause MD in at least 1 per 5,000 individuals.^4,5^ To date, 127 high-confidence “confirmed” pathogenic SNV/indel variants have been reported in the expert-curated database MITOMAP.^6^ The majority of these variants cause disease in the heteroplasmic state, when the heteroplasmy level (HL) of mtDNA molecules carrying the variant exceeds a critical threshold in a susceptible tissue, typically reported as 60-80%.^7^ mtDNA heteroplasmy increases the complexity of genetic diagnosis, as HL can vary from tissue to tissue. HL is typically highest in post-mitotic tissues such as skeletal muscle, heart, and brain, and is often lowest in rapidly replicating non-disease affected tissues that are more readily accessible to sampling and routinely used first-line for DNA testing, in particular blood and buccal cells.^8^ In comparison, only a small number of homoplasmic variants have been associated with disease. These variants often demonstrate incomplete penetrance whereby only a subset of variant carriers manifest with the disease, as is commonly reported for the m.11778G>A variant causing Leber Hereditary Optic Neuropathy (LHON) or lead to adult-onset and milder disease.^9^

Distinguishing mtDNA-encoded disease from other mitochondrial and non-mitochondrial nuclear-encoded diseases is clinically challenging due to the phenotypic heterogeneity of MDs and overlap with other nuclear-encoded neurological, neuromuscular, ophthalmological, and hematological diseases,^1,10^ among others. It is, however, essential in determining the mode of inheritance to inform genetic counseling, provide accurate recurrence risk estimates, and can be important in disease prevention, such as by egg donation, mitochondrial transfer, or preimplantation genetic diagnostics,^11^ as well as to implement preventative measures and anticipatory care.

To reduce sequencing cost and to streamline data analysis in rare disease diagnostics, the mtDNA does not routinely undergo targeted sequencing and analysis unless a MD is clinically suspected. This potentially leads to cases of mtDNA-encoded MDs eluding detection when exome (ES) or genome (GS) sequencing are selected as the first-line diagnostic test.^12^ Analysis of the mtDNA is, nevertheless, possible in a holistic approach from ES^13,14^ and GS data^15^ by applying dedicated bioinformatic pipelines to call mtDNA SNVs/indels,^16,17^ and large mtDNA deletions.^18^ For ES, probes can be added to the library preparation to capture the mtDNA at high coverage^19^. Alternatively, off-target reads can be analyzed, though this approach provides relatively low coverage of the mtDNA and is more likely to be enriched for nuclear DNA of mitochondrial origin (NUMTs).^14^ In comparison, GS provides high coverage of the mtDNA due to the naturally high copy number of mtDNA molecules in cells.^15,16^ mtDNA-specific bioinformatic pipelines navigate alignment issues created by the circular nature of the mtDNA, facilitate the detection of variants at low heteroplasmy level (not possible in routine variant calling pipelines), and apply strategies to reduce the misalignment of NUMTs that can otherwise result in false positive putative heteroplasmies. These pipelines have proven successful in the diagnosis of mtDNA encoded disease in cohorts of suspected mitochondrial and neurological disease.^13–15,20^

Here, we apply mtDNA variant calling pipelines to GS, ES, and, where available, RNA-sequencing data from a diverse collection of over 6,500 rare disease families primarily sequenced through the GREGoR (Genomics Research to Elucidate the Genetics of Rare diseases) Consortium. We search for reported pathogenic variants and leverage recently released reference population databases of homoplasmic and heteroplasmic mtDNA variant allele frequencies (gnomAD v3^16^ and HelixMTdb^21^) in combination with mtDNA-specific computational prediction tools and mitochondrial constraint metrics^22^ for novel variant prioritization and assessment.

## METHODS

### Sample selection

ES, GS, and RNA-sequencing data (when available) from probands with a suspected rare disease and their affected and unaffected family members, recruited, sequenced, and phenotyped by the GREGoR Consortium (U07), were subject to mtDNA variant calling. In addition, samples from the Broad Institute Center for Mendelian Genomics (Broad CMG) that could not be part of GREGoR due to disease-specific consent, along with other rare disease cohorts sequenced in collaboration with the Broad CMG were included in the analysis. This resulted in a total of 14,282 samples from 7,282 families. ES libraries were generated using either Nextera capture (no mtDNA probes included) or Twist capture (with mtDNA probes included), and all samples were sequenced using Illumina instruments. Samples with a high level of contamination (≥2% of haplogroup defining variants at 85-99.8% heteroplasmy level) and/or a mean per sample mtDNA coverage of <20X were excluded from the study, resulting in 13,160 samples from 6,660 families for mtDNA variant analysis (Fig. S1).

### mtDNA variant calling, haplogroup determination, and variant annotation

mtDNA variants were called from GS data using the mitochondria mode of GATK- Mutect2^16^ and from ES/RNA-sequencing using the MToolBox pipeline.^17^ RNA was processed using a stranded polyA-tailed kit (Illumina). Large mtDNA deletions were called using MitoSAlt.^18^ Variants were annotated with quality flags, functional consequence, reference population frequency, computational predictions, and mitochondrial constraint metrics.^22^ Variants flagged as low quality were removed (see Supplemental Methods for more details).

### Identifying pathogenic mtDNA variants

Variants with “confirmed” disease-causing status were extracted from MITOMAP (n=127, last accessed October 2024).^6^ Variants submitted as P/LP with ≥2-star review status in association with primary MD were extracted from ClinVar (n=111, last accessed October 2024).^23^ This resulted in a total of 152 unique reported P/LP variants for analysis (Table S1).

### Identifying high priority novel variants

Novel variants were filtered to: i) non-haplogroup defining variants (for the haplogroup of the respective sample), ii) non-synonymous variants; iii) rare variants detected in <1:50,000 individuals at homoplasmy in reference populations (gnomAD v3 and HelixMTdb) and with an allele count ≤10 across all samples in the call set, and iv) variants meeting at least one of the following criteria for predicted deleteriousness: i) predicted loss-of-function (frameshift, stop gained), ii) missense with an APOGEE2 score >0.5^24^ and/or HmtVar score ≥0.35^25^), iii) tRNA with MitoTIP score >12.66^26^, PON-mt-tRNA probability score ≥0.5^27^, and/or HmtVar score ≥0.35; iv) within an area of regional constraint or at a nucleotide position with high mitochondrial local constraint (MLC score ≥0.75)^22^ (see Supplemental Methods for more details).

### Variant interpretation and confirmation

Identified variants were clinically evaluated as either: i) Diagnostic: A previously reported or novel variant classified as P/LP according to the ClinGen Variant Curation Expert Panel mtDNA-specifications of the American College of Medical Genetics and Genomics and Association of Molecular Pathologists (ACMG/AMP) standards and guidelines for variant interpretation,^28^ that explains the proband’s phenotype, is detected at a clinically relevant HL, for which the multidisciplinary analysis team and referring clinician consider the variant causative, and clinically confirmed in a CLIA certified laboratory; ii) Candidate: A reported P/LP variant or high priority variant of uncertain significance (VUS) that may explain the proband’s phenotype but requires additional evidence to establish causality and/or pathogenicity; iii) Pathogenic variant of undetermined clinical relevance: A reported P/LP variant that does not explain the individual’s phenotype and/or is known to demonstrate incomplete penetrance at near-homoplasmy.

### Phenotype data analyses

Phenotype data were collected as Human Phenotype Ontology (HPO) terms. Each reported HPO term was mapped in the ontology to “phenotypic abnormality” (HP:0000118) and annotated with all intermediate terms. The objective clinical likelihood of the proband having a disease of mitochondrial etiology was calculated by the Mitochondrial Disease Criteria (MDC) score,^29^ adapted for use with HPO terms^10^. MDC scores were stratified into unlikely (score 0-1), possible (score 2-4), probable (score 5-7), and definite (score 8-12) MD.

## RESULTS

### Cohort description

In total, 6,660 families were included in our analysis after sample-level quality filtering (see **Methods**). The majority (5,625, 84%) were genetically undiagnosed following nuclear analysis of ES/GS (**Fig. 1a**). Among the solved families were three that had already had an mtDNA-encoded diagnosis returned by targeted mtDNA sequencing, used as positive controls for our variant calling and analysis pipelines. Data from multiple sequencing methods (ES, GS, and/or RNA-sequencing) were analyzed for 164 probands (**Fig. 1b**).

**Fig 1.**
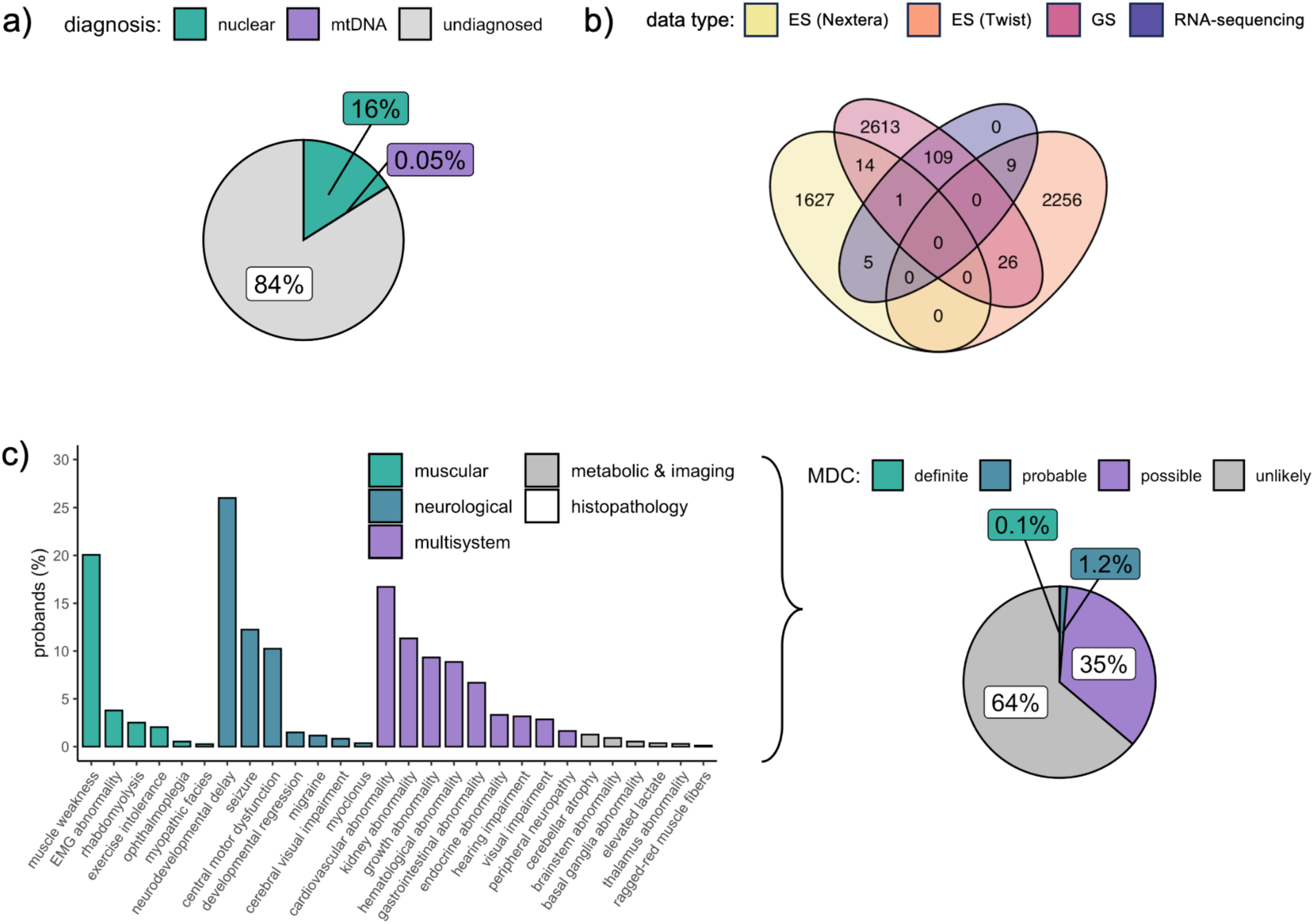
Study cohort overview. **a)** Diagnostic status of probands following nuclear variant analysis by ES and/or GS, prior to the analysis of mtDNA variants in this study. For three probands (0.05%) diagnostic variants in the mtDNA had already been detected by targeted mtDNA sequencing. **b)** Proband samples by sequencing type, demonstrating overlap in data type for analysis. **c)** Frequency of HPO terms indicative of MD in the probands (displaying terms reported in ≥5 probands) and the resultant MDC classification of clinical likelihood of a MD.^29^

Most samples were derived from DNA extracted from blood from probands with pediatric onset of disease, and therefore less likely to carry mtDNA variants restricted to post-mitotic tissues. A median of three non-redundant HPO terms were reported per proband (range 0-121). For 5,192 probands (78%), at least one reported HPO term overlapped with a term associated with MDs, according to the Mitochondrial Disease Criteria (MDC),^29^ spanning muscular, neurological, multisystem, metabolic, imaging, and histopathology terms (**Fig. 1c**). Based on the combination of these phenotypes and applying the MDC, 1.2% of probands had a probable or definite likelihood of a MD, 35% possible, and 64% unlikely, prior to genetic analysis. These figures indicate a low prior probability of a mitochondrial disease based upon clinical phenotype for most families in our study.

### mtDNA coverage and variant detection summary

The mean per-base mtDNA coverage for GS and RNA-sequencing was high (GS mean 4,416x, RNA-sequencing mean 5,894x). The coverage by ES depended on capture selection. ES (Twist) provided high coverage (mean 6,315x) by adding mtDNA probes, whereas ES (Nextera) provided low coverage from off-target reads (mean 47x) (**Fig. 2**).

**Fig 2.**
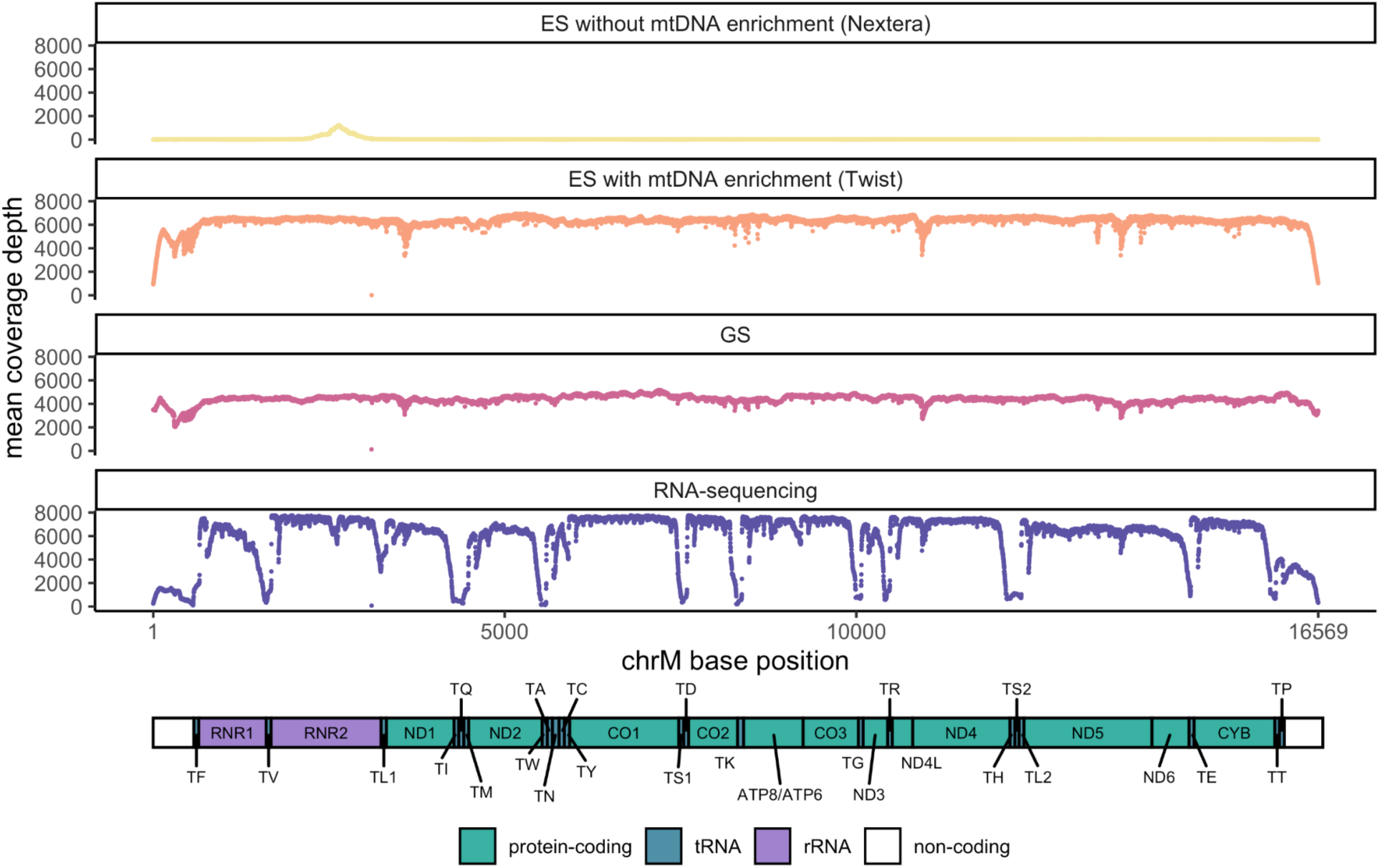
mtDNA coverage by data type. Mean per-base mtDNA coverage by chrM base position. Schematic of a linearized mtDNA, demonstrating lowest coverage over the artificial break in the mitochondrial D-loop and low RNA-sequencing coverage of the tRNA genes due to lack of a stable 3’-poly(A) tail for enrichment ^30^.

A mean of 40 mtDNA variants were called at ≥1% HL per sample, of which 26 per sample passed our quality filters (see Supplementary Methods). Collectively, 6,960 unique variants were detected, spanning 6,069 of the 16,569 nucleotide positions of the mtDNA (37%). Most were detected at near-homoplasmy (≥95% heteroplasmy level) and were known haplogroup defining variants (mean 23 per sample), that are unlikely to be causal of MDs. A summary of the counts of high-quality variants per proband sample for analysis by data type is displayed in Table S2.

Maternal samples were available for 3,056 probands (46%), allowing comparison of heteroplasmy level between generations that can be informative for clinical interpretation. Overall, 99% (80,753/81,772) of variants in the probands were detected in the maternal sample of the corresponding data type, >99% of homoplasmic variants (77,449/77,767) and 82% of heteroplasmic variants (3,304/4,005). A small number of these variants demonstrated either a potentially clinically relevant positive heteroplasmic shift, from below to above the “typical” disease-causing threshold of 60% (147/80,753, 0.18%) or negative heteroplasmic shift, from above to below 60% heteroplasmy level (85/80,753, 0.11%), though the majority were more neutral (Fig. S2). Variants detected in the proband only (1,019/81,772 variants, 1.2%) may be *de novo*, somatic, or present at undetectable levels in the maternal tissue sampled. Over one-third of the variants detected in the proband-only were at a heteroplasmy level ≥60% (388/1,019, 38%) and, when predicted to be deleterious, are promising candidates for sporadic disease in the proband.

### Detection of reported P/LP variants

Reported P/LP variants were detected in a total of 59 probands. Large mtDNA deletions were detected in two of these probands (**Fig. 3a**) and pathogenic mtDNA SNVs or small indels, reported in MITOMAP as “confirmed” disease-causing and/or reported in ClinVar as P/LP with ≥2-star review status, were identified in the remaining 57 probands (24 different variants) at ≥5% HL (**Fig. 3b**). In total, nine new diagnoses were made (including one large deletion), two plausible candidate diagnoses were identified, and all three of the known mtDNA diagnoses in the cohort (including one large deletion) were reidentified (**Table 1**). In the remaining 45 probands, the pathogenic variants were of undetermined clinical relevance.

**Fig 3.**
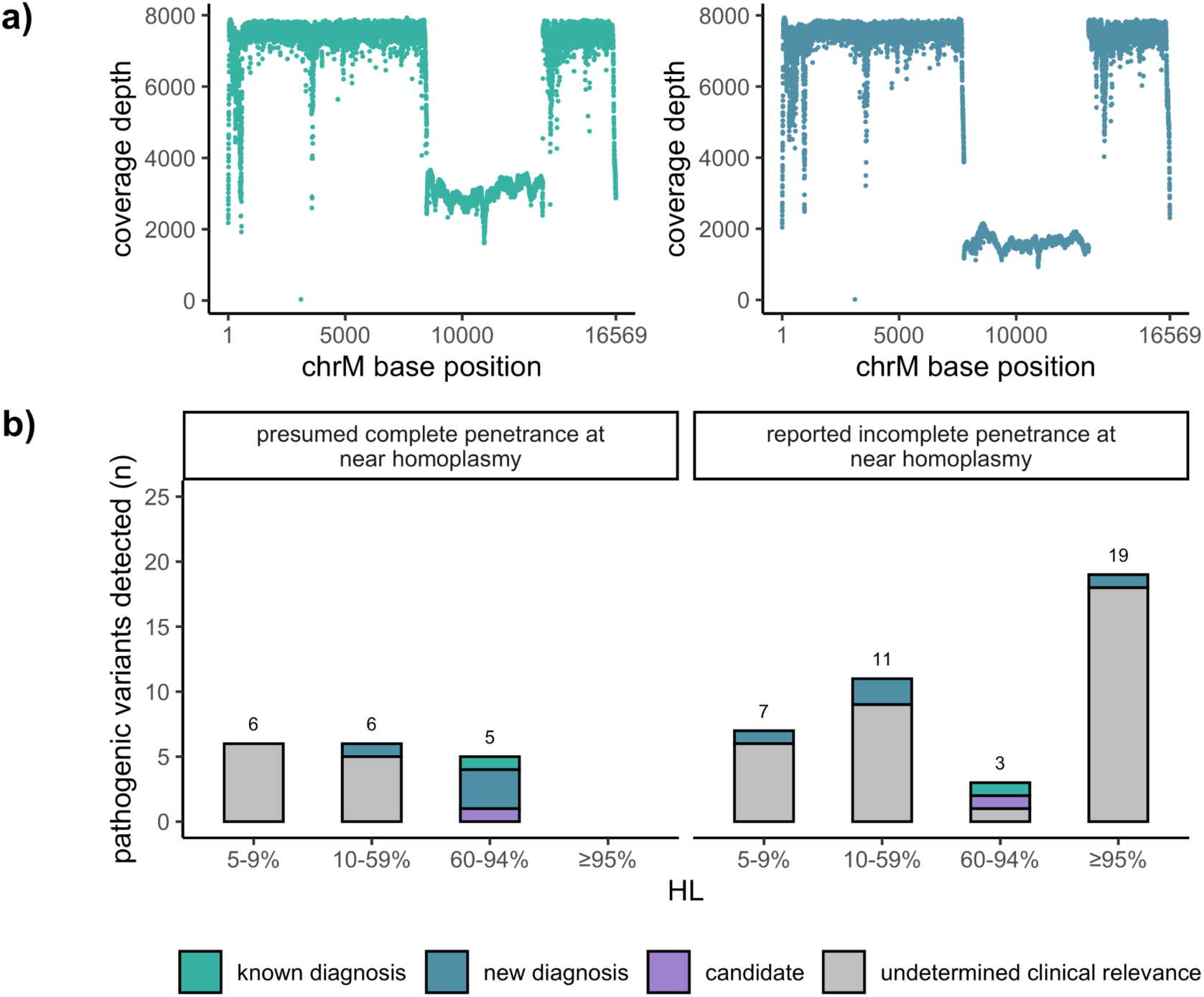
Detection of pathogenic mtDNA variants. **a)** Coverage by chrM base position in two samples with large mtDNA deletions (green [known diagnosis] and blue [new diagnosis]). **b)** Pathogenic variants detected across all probands, stratified by penetrance at near homoplasmy.^13^

**Table 1.** Diagnostic and candidate reported pathogenic mtDNA variants.

| Proband ID | Sample data type | Gene symbol | Variant (consequence) | MITOMAP status | ClinVar status | Sample HL (alt/ref reads) | Inheritance | Reported phenotype | Proband's Phenotype | Proband's MDC (score) | Diagnostic outcome |
| --- | --- | --- | --- | --- | --- | --- | --- | --- | --- | --- | --- |
| P1 | ES | *MT-ND1* | m.3946G>A (missense) | rptd | P/LP | 70% (2,924/1,247) | - | MELAS | Infantile spasms, cortical visual impairment, and severe NDD | Possible (3) | Known diagnosis |
|  | RNA-seq |  |  |  |  | 68% (4,720/2,231) |  |  |  |  |  |
| P2 | ES | *MT-ATP6* | m.8993T>G (missense) | cfrm | P | 87% (5,473/847) | - | NARP, LSS, MILS, LHON | Retinitis pigmentosa | Unlikely (1) | Known diagnosis |
| P3 | ES | - | m.8470_13446del4977 (single deletion) | - | - | 70% (4,701/1,958) | - | Pearson syndrome | Pearson syndrome and DBA | Possible (3) | Known diagnosis |
| P4 | ES | - | m.7773_13094del4977 (single deletion) | - | - | 82% (5904/1316) | - | Pearson syndrome | Pearson syndrome and CSA | Possible (4) | New diagnosis |
| P5 | ES | *MT-TL1* | m.3243A>G (tRNA) | cfrm | P/LP | 8% (490/5,634) | - | MELAS, LSS, MIDD, SNHL, CPEO, myopathy, FSGS, and ASD | Retinitis pigmentosa | Unlikely (1) | New diagnosis |
| P6 | GS | *MT-TL1* | m.3243A>G (tRNA) | cfrm | P/LP | 10% (501/5,014) | - | MELAS, LSS, MIDD, SNHL, CPEO, myopathy, FSGS, and ASD | Fatigue, periodic fevers, headache, recurrent fever, tinnitus, recurrent episodes of infection, intermittent rashes, significant nausea | Possible (2) | New diagnosis |
| P7 | ES | *MT-TL1* | m.3243A>G (tRNA) | cfrm | P/LP | 11% (484/3,959) | - | MELAS, LSS, MIDD, SNHL, CPEO, myopathy, FSGS, and ASD | Retinitis pigmentosa | Unlikely (1) | New diagnosis |
| P8 | ES | *MT-TL1* | m.3243A>T (tRNA) | cfrm | P/LP | 33% (1,384/4,194) | Transmitted (1.4% HL in maternal sample) | Myopathy, MELAS, SNHL, CPEO | Mitochondrial myopathy (RRF, COX- fibers), fatigue, growth delay | Probable (7) | New diagnosis |
| P9 | GS | *MT-TA* | m.5591G>A (tRNA) | rptd | P | 86% (4,253/710) | - | Myopathy | Mitochondrial myopathy (RRF, COX- fibers), exercise intolerance, elevated CK, headaches, ID, tachycardia | Definite (8) | New diagnosis |
| P10 | GS | *MT-ATP6* | m.8969G>A (missense) | cfrm | LP | 70% (3,646/1,540) | - | MLASA, IgG nephropathy | LSS, hypotonia, FTT, episodic vomiting | Probable (6) | New diagnosis |
| P11 | ES | *MT-ATP6* | m.8993T>C (missense) | cfrm | P | 99% (7,018/71) | Transmitted (90% HL in maternal sample) | NARP, LSS, MILS, LHON | Hypotonia, cerebellar atrophy, speech delay | Possible (3) | New diagnosis |
| P12 | GS | *MT-ATP6* | m.9134A>G (missense) | rptd | LP | 82% (4,921/6,038) | *de novo* | IUGR, hypotonia, HCM, and LA | SGA, LA, NDD, infantile spasms | Probable (5) | New diagnosis |
| P13 | GS | *MT-TF* | m.591C>T (tRNA) | rptd | P/LP | 72% (4,388/963) | Transmitted (36% HL in maternal sample) | Renal tubulopathy | Renal tubulopathy, ID, NDD, short stature | Possible (4) | Candidate |
| P14 | GS | *MT-TL1* | m.3243A>T (tRNA) | cfrm | P/LP | 70% (3,373/4,819) | Transmitted (9% HL in maternal sample) | Myopathy, MELAS, SNHL, CPEO | SNHL, cerebral palsy, holoprosencephaly* | Possible (2) | Candidate |

cfrm, confirmed; rptd, reported; P, pathogenic; LP, likely pathogenic; HL, heteroplasmy level, DBA, Diamond Blackfan anemia; MLASA, Mitochondrial myopathy, lactic acidosis and sideroblastic anemia; LSS, Leigh syndrome spectrum; MELAS, Mitochondrial Encephalopathy, Lactic Acidosis, and Stroke-like episodes; NDD, neurodevelopmental delay; CPEO, Chronic progressive external ophthalmoplegia; NARP, Neuropathy, Ataxia, Retinitis Pigmentosa; HCM, hypertrophic cardiomyopathy; CSF, cerebrospinal fluid; MIDD; Maternally inherited diabetes and deafness; SNHL, sensorineural hearing loss; FSGS, focal segmental glomerulosclerosis; ASD, autism spectrum disorder; CSA, congenital sideroblastic anemia; CMS; congenital myasthenic syndrome; LA, lactic acidosis; FTT, failure to thrive; EM, encephalomyopathy; LHON, Leber Hereditary Optic Neuropathy; IUGR, intrauterine growth restriction; OXPHOS, oxidative phosphorylation; CLIA, Clinical Laboratory Improvement Amendments. \*LP variant in *ZIC2* explains the proband's holoprosencephaly phenotype, *MT-TL1* is a candidate for the SNHL and would be a dual diagnosis.

Non-diagnostic pathogenic variants may be detected in individuals, including those in reference populations, at a heteroplasmy level below the disease-causing threshold (typically reported at ≥60%, though dependent on the specific variant and tissue) and at near homoplasmy when the variant demonstrates incomplete penetrance or is associated with adult-onset or mild disease.^13,31^ Stratifying the detected pathogenic variants by reported incomplete penetrance at near homoplasmy, we find all P/LP variants at high heteroplasmy level, without reports of incomplete penetrance to be diagnostic, with the proband’s phenotype being in-keeping with reported phenotypes for the variant (**Figure 3b**). In contrast, we detected many non-diagnostic P/LP variants of undetermined clinical relevance at high heteroplasmy level that are reported to be incompletely penetrant.

The most frequently detected non-diagnostic pathogenic variants were: i) m.1555A>G in *MT-RNR1* (10 probands) associated with susceptibility to aminoglycoside ototoxicity, ii) m.3243A>G in *MT-TL1* (10 probands) associated with Mitochondrial Encephalopathy, Lactic Acidosis, and Stroke-like episodes (MELAS), though highly phenotypically heterogeneous; iii) m.11778G>A in *MT-ND4* (7 probands) associated with Leber Hereditary Optic Neuropathy (LHON). Both m.1555A>G and m.11778G>A are known to demonstrate incomplete penetrance at near-homoplasmy.^13^ There are also numerous reports of asymptomatic individuals with m.3243A>G at high heteroplasmy level in blood (≥60%).^32^

### Detection of novel variants

We next sought to investigate whether novel mtDNA variants may be causing disease. To prioritize variants with high disease-causing potential, we applied stringent filtering by function, frequency in reference populations, predicted deleteriousness, and mitochondrial constraint metrics (see **Methods**). In total, 555 variants were prioritized in 518 probands (0.08 per proband across all analyzed probands) (**Fig. 4**). Each variant was carefully reviewed for potential relevance to phenotype and, if considered of diagnostic interest, was classified according to the ACMG/AMP standards and guidelines for mitochondrial DNA variant interpretation. This clinical evaluation led to one new diagnosis and one candidate diagnosis classified as Likely Pathogenic (LP) by the mtDNA-specifications of the ACMG/AMP,^28^ in addition to eight high-priority candidates classified as VUS (**Table 2**).

**Fig 4.**
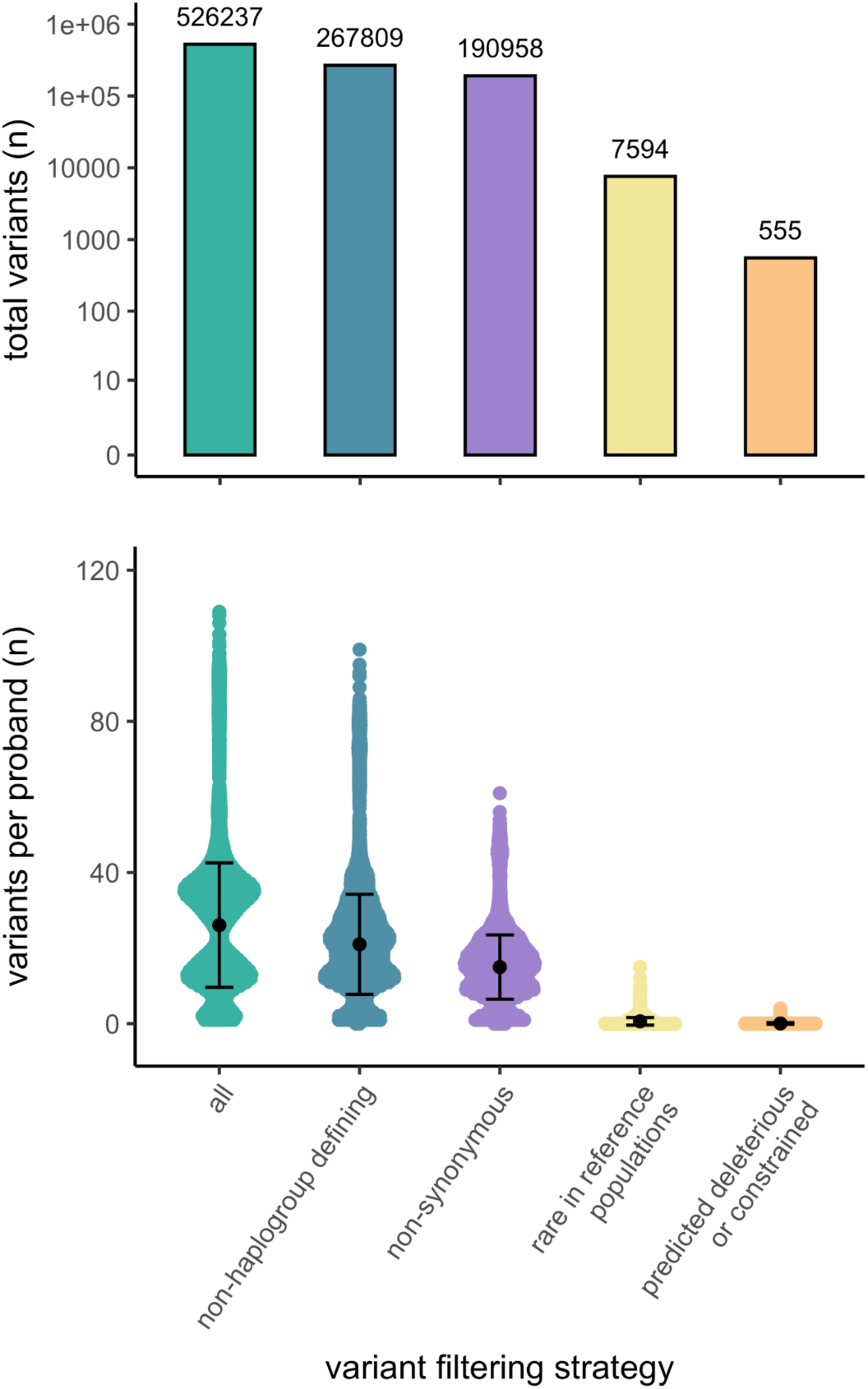
Prioritization of novel variants for clinical evaluation. Number of variants in total (top panel) and per proband (bottom panel) for clinical evaluation, displayed with the mean and s.d.

**Table 2.** Diagnostic and candidate novel mtDNA variants.

| Proband ID | Sample data type | Gene symbol | Variant (consequence) | MITOMAP status | ClinVar status | Sample HL (alt/ref reads) | Heteroplasmic AF (AC, max observed HL) | Computational evidence of deleteriousness | Mitochondrial constraint | Transmission | Reported phenotype | Proband's Phenotype | Proband's MDC (score) | ACMG classification | Diagnostic outcome |
| --- | --- | --- | --- | --- | --- | --- | --- | --- | --- | --- | --- | --- | --- | --- | --- |
| P15 | ES | MT-CYB | m.15347C>T (missense) | - | - | 19% (7/30) | gnomAD: absent<br>Helix: absent | APOGEE 0.64, HmtVar 0.84 | Regional missense constraint MLC:0.53 | *de novo* | - | HMC, LA, elevated CSF lactate | Probable (6) | LP | New diagnosis |
| P16 | GS | MT-TP | m.16023G>A (tRNA) | rptd | VUS | 15% (1,584/8,977) | gnomAD: absent<br>Helix: 1.53e-05 (3, 15%) | MitoTIP 17.6, HmtVar 0.65, PON-mt-tRNA 0.8 | MLC:0.73 | *de novo* | Migraine, pigmentary retinopathy, deafness, leukoaraiosis | Seizures, FTT, neutropenia, anemia | Possible (3) | LP | Candidate |
| P17 | GS | MT-CO1 | m.6853G>C (missense) | - | - | 6% (308/4,829) | gnomAD: absent<br>Helix: absent | APOGEE 0.52, HmtVar 0.8 | MLC:0.87 | Transmitted (2% HL in maternal sample) | - | Seizures, speech and language delay, myopathy | Possible (4) | VUS | Candidate |
| P18 | GS | MT-TK | m.8328G>A (tRNA) | rpt | VUS | 6% (537/8,952) | gnomAD: absent<br>Helix: 5.10e-06 (1, 14%) | MitoTIP 117.5, PON-mt-tRNA 0.92 | MLC:0.62 | *de novo* | Encephalopathy / EXIT with myopathy and ptosis | Hypotonia, stridor, feeding difficulties, laryngomalacia, chronic constipation, fatigue, disturbed sleep, aerophagia, reflux, asthma, periodic limb movement, muscle fatigue, loss of skill, plagiocephaly, neck flexion weakness | Possible (3) | VUS | Candidate |
| P19 | GS | MT-ATP8 | m.8424T>C (missense) | rptd | - | 96% (2,265/2,359) | gnomAD: absent Helix: 5.10e-06 (1, 15%) | APOGEE2 0.55, HmtVar 0.85 | MLC:0.01 | *de novo* | - | LSS, FTT, hypotonia, seizures, regression | Probable (5) | VUS | Candidate |
|  | RNA-seq |  |  |  |  | 98% (7,560/195) |  |  |  |  |  |  |  |  |  |
| P20 | ES | MT-ATP8 | m.8570T>C (stop-loss) | - | - | 95% (6,868/7,229) | gnomAD: 3.54e-05 (2, 28%) Helix: absent | LoF | MLC:0.03 | - | - | CSA | Possible (4) | - | Candidate |
| P21 | ES | MT-ATP6 | m.8611C>A (missense) | - | - | 100% (5,651/0) | gnomAD: absent Helix: absent | APOGEE 0.45, HmtVar 0.71 | MLC:0.01 | - | - | NDD, cerebellar atrophy, strabismus | Possible (3) | VUS | Candidate |
| P22 | ES | MT-ATP6 | m.8797T>C (missense) |  |  | 50% (3,694/7,387) | gnomAD: absent Helix: absent | APOGEE 0.39 HmtVar 0.85 | MLC:0.15 | - | - | LSS | Possible (3) |  | Candidate |
| P23 | ES | MT-TH | m.12197C>T (tRNA) | - | - | 100% (7,776) | gnomAD: 1.77e-05 (1, 17%) Helix: 1.02e-05 (2, 29%) | MitoTIP 14.6, PON-mt-tRNA 0.62 | MLC:0.27 | transmitted (57% HL in maternal sample) | - | CSA, SNHL, ID, motor delay | Possible (4) | VUS | Candidate |
| P24 | ES | MT-TH | m.12198T>C (tRNA) | - | - | 87% (6,707/1,002) | gnomAD: absent Helix: 5.10e-06 (1, 9%) | MitoTIP 17.9, PON-mt-tRNA 0.53 | MLC:0.28 | - | - | CSA | Possible (4) | VUS | Candidate |

HL, heteroplasmy level; DBA, Diamond Blackfan anemia; MLASA, Mitochondrial myopathy, lactic acidosis and sideroblastic anemia; LSS, Leigh syndrome spectrum; MELAS, Mitochondrial Encephalopathy, Lactic Acidosis, and Stroke-like episodes; NDD, neurodevelopmental delay; CPEO, Chronic progressive external ophthalmoplegia; RP, Retinitis pigmentosa; NARP, Neuropathy, Ataxia, Retinitis Pigmentosa; HCM, hypertrophic cardiomyopathy; CSF, cerebrospinal fluid; MIDD, Maternally inherited diabetes and deafness; SNHL, sensorineural hearing loss; FSGS, focal segmental glomerulosclerosis; ASD, autism spectrum disorder; CSA, congenital sideroblastic anemia; CMS, congenital myasthenic syndrome; LA, lactic acidosis; FTT, failure to thrive; EM, encephalomyopathy

The novel LP diagnosis is a *de novo* m.15347C>T (p.His201Tyr) variant in *MT-CYB*, detected at 19% heteroplasmy level in the blood of a genetically undiagnosed proband (P15) who presented in the neonatal period with progressive hypertrophic cardiomyopathy, renal cortical dysplasia, hyperinsulinemic hypoglycemia, and elevated lactate in both the serum and cerebrospinal fluid. *MT-CYB* encodes a subunit of mitochondrial complex III. The p.His201 amino acid position has high conservation (MITOMASTER 100% across species) and is in an area of regional missense constraint,^22^ with this residue thought to be critical for ubiquinone binding^33^. The variant has consistently deleterious computational predictions (APOGEE2 0.64, HmtVar 0.84), is absent in reference populations at both homo- and heteroplasmy (gnomAD v3 and HelixMTdb), and has not previously been reported in clinical cases. Follow-up by targeted whole mitochondrial genome analysis on DNA extracted from heart tissue found the variant to be present at 87.5% HL. The variant was initially classified as a VUS. Subsequent segregation testing of maternal DNA extracted from both blood and urine was negative, suggesting the variant to be *de novo*. Functional studies were also performed, demonstrating the variant to have a deleterious effect on levels of the MT-CYB protein and on complex III activity and protein levels in affected tissues (heart and muscle, data not shown). With these additional lines of evidence for pathogenicity, the variant was reclassified as LP (PS2, PS3_Supporting, PM2_Supporting, PP3_Supporting) and was returned to the family to inform family planning.

The novel LP candidate is a *de novo* m.16023G>A variant in *MT-TP*, detected at 15% heteroplasmy level in blood in a proband (P16) presenting in infancy with unexplained seizures, failure to thrive, pancreatic exocrine insufficiency, neutropenia, anemia, lethargy, and recurrent infections typically with hemodynamic instability requiring intensive care admission. The variant was absent in the mother’s GS data. *MT-TP* encodes a mitochondrial tRNA. The variant has consistently deleterious computational predictions (MitoTIP 17.6, HmtVar 0.65, PON-mt-tRNA 0.8). It is absent in reference populations at homoplasmy and is rare at heteroplasmy (absent in gnomAD v3, heteroplasmic allele count of three in HelixMTdb with a maximum detected HL of 15%). The variant is listed in MITOMAP with “reported” status, and was previously reported in two unrelated probands. In the first proband with migraine, pigmentary retinopathy, deafness, leukariosis on magnetic resonance imaging (MRI), COX-negative fibers and ragged red fibers on muscle biopsy (proband HL 9% in blood, 86% in muscle, and 36% in urine; mother HL 1% in blood, 7% in urine) the variant is reported to be diagnostic. In the second proband with liver dysfunction, urticaria, developmental delay, and fatigue (HL 2% in muscle), the variant remains of undetermined clinical relevance.^34,35^ The variant has been functionally validated by gold-standard single fiber analysis.^34^ Given these lines of evidence, the variant is classified as LP (PM6, PS3_Supporting, PS4_Supporting, PM2_Supporting, PP3_Supporting). Clinical follow-up is underway to measure the variant’s HL in additional tissues from the proband and mother to determine if this is diagnostic for the family. The remaining eight high-priority candidates are reported in **Table 2**.

### Detection of excess heteroplasmic variants provides functional evidence of pathogenicity for a *de novo* variant in the proofreading exonuclease domain of *POLG*

As part of our analysis, we counted the number of heteroplasmic variant calls per proband at ≥1% heteroplasmy level. This analysis revealed an outlier proband (P25) in the ES (Twist) data set with 941 heteroplasmic variants (910 SNVs, 31 indels), compared to a mean of 10.1 per sample (**Fig. 5a**). Only 12 of the 941 variants were detected in the corresponding maternal sample, indicating either an issue with sample quality (despite passing our sample-level coverage and contamination filters) or pointing towards an underlying defect in the replication and repair of the mtDNA leading to high rates of somatic SNVs. Analysis of ES from the proband and both unaffected parents for causal nuclear variants had detected a novel *de novo* missense variant that was initially of uncertain significance in the nuclear gene *POLG*, encoding DNA polymerase gamma, (c.592G>A, p.Asp198Asn), responsible for mtDNA replication and repair. This variant affects the p.Asp198 residue, an essential catalytic residue involved in the POLG protein’s proofreading exonuclease activity, and is predicted to be deleterious (REVEL score 0.94) (**Fig. 5b**). In reported cellular models, mutagenesis of p.Asp198 to p.Asp198Ala abolishes the exonuclease activity of POLG^36^ and, in keeping with the finding of a high heteroplasmic variant detection rate in our proband, results in accumulation of somatic SNVs in the mtDNA.^37^ Similarly, the *POLG* mutator mouse that is lacking the mtDNA proofreading exonuclease activity rapidly accumulates somatic mtDNA SNVs.^38^ The proband’s p.Asp198Asn *POLG* variant is absent in reference population databases. The proband presents with congenital sideroblastic anemia (CSA), leukopenia, moderate neutropenia, and lymphopenia, with no associated history of recurrent infections. Short stature (1st centile) as well as cognitive and learning disabilities are also reported, and the brain MRI demonstrates polymicrogyria. Early type I diabetes mellitus was also recently diagnosed. A CSA phenotype is reported only once in association with a heterozygous *POLG* variant in the literature,^39^ yet is highly consistent with both nuclear- and mtDNA-encoded MD.^40^ Moreover, among the many heteroplasmic mtDNA variants detected in the proband was a rare, somatic, predicted deleterious (APOGEE2 0.76) missense variant in *MT-ND1* (m.3976T>C, p.Phe224Leu). The p.Phe224 amino acid position has high conservation (MITOMASTER 97.8% across species) and is in an area of regional missense constraint, and the nucleotide position (m.3976T) has high mitochondrial local constraint (MLC score 0.86). *MT-ND1* encodes a subunit of mitochondrial complex I. The variant is detected at 21% HL and may explain the hematological manifestation of disease in this proband (**Fig. 5c**). For the two GS samples with excess heteroplasmic variants, no potentially causal rare variants were identified in genes involved in mtDNA replication and repair.

**Fig. 5.**
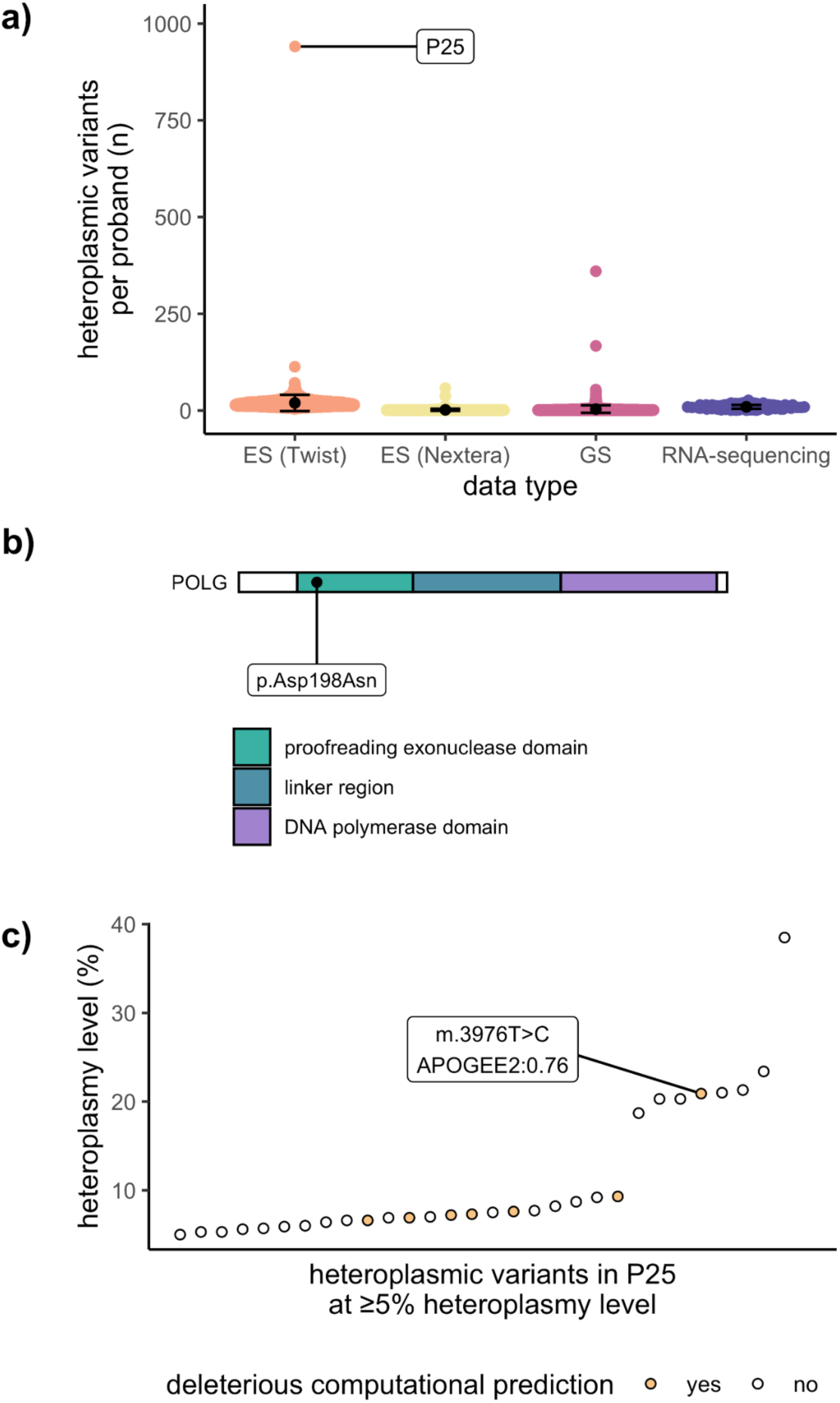
Novel *de novo* variant in *POLG* results in high numbers of somatic heteroplasmic mtDNA variants. a) Number of high-quality mtDNA variants per proband by data type, demonstrating P25 as an outlier sample with >900 variants. b) Schematic representation of POLG. The *de novo POLG* variant in P25 is located within the proofreading exonuclease domain of the protein. c) Heteroplasmic mtDNA variants detected in P25 at ≥5% HL indicating those with deleterious computational prediction.

### Diagnostic detection rate

In summary, across the 6,660 probands included in our analysis, a total of 614 variants were prioritized for clinical evaluation, spanning reported pathogenic SNV and indel variants (57 total), large mtDNA deletions (2 total), and prioritized novel variants (555 total) - approximately one variant per 10 probands. Our variant calling and analysis pipelines reidentified all three known mtDNA diagnoses in the previously solved families with targeted mtDNA sequencing (including one large mtDNA deletion) and established 10 new diagnoses among the undiagnosed families. Candidate diagnoses were also identified in 11 probands from undiagnosed families that remain under investigation (e.g., sequencing of additional tissues, sequencing additional maternal family members). Together with the novel *POLG* diagnosis, findings from the analysis of the mtDNA resulted in a diagnostic uplift of 0.4% (22/5,625) in undiagnosed families with a generally low prior probability of a MD.

## DISCUSSION

We evaluated the diagnostic yield of calling mtDNA variants from ES and GS data that had already been generated and analyzed for suspected Mendelian (nuclear) causes of disease. This followed the rationale that mtDNA-encoded MDs may be overlooked in the differential diagnoses when referring a family for genetic testing, due to extensive overlap with seemingly Mendelian phenotypes, including nuclear causes of MDs that are more frequent in children.

We provided a diagnosis to a total of 11 previously undiagnosed families, in addition to identifying candidates in a further 11, presenting clinically with a broad range of rare diseases. Many of our candidates were identified in probands with congenital sideroblastic anemia, a phenotype with a very high probability of being an MD, either associated with nuclear-encoded mitochondrial proteins or encoded by the mtDNA.^40^ According to our calculation of the MDC based upon available HPO terms, many candidates were also detected in probands with phenotypes indicating an “unlikely” or “possible” likelihood of MD including retinal disease, a phenotype for which nuclear variants are more frequently investigated.^40,41^ Our diagnostic findings demonstrate the value of adding mtDNA analysis to routine ES/GS data analysis for probands where a nuclear-encoded mitochondrial cause is considered likely, as well as for those with a low prior-probability of MD. Moreover, for all probands with maternal data available, the diagnostic or candidate variants were found to either arise *de novo*, as reported for ∼20% of mtDNA-encoded disease,^42^ or to have been transmitted from the unaffected mother with an increase in heteroplasmy level, presumably at the point of the “mitochondrial bottleneck” in development.^38^ Therefore, the absence of a maternal family history should not exclude the suspicion of an mtDNA-encoded disease. The diagnostic uplift of 0.4% among undiagnosed families was in line with expectation based on published studies demonstrating diagnostic rates of 0.1% (11/11,424)^14^ in individuals enriched for neurological diseases and 1.6% (5/319)^15^ to 1.8% (38/2,111)^13^ in individuals with higher clinical suspicion of MD.

We also report on the overall rate of pathogenic mtDNA variant detection among sequenced probands by heteroplasmy level threshold, finding P/LP variants in many probands to be secondary findings for conditions/genes not included on the ACMG recommended reporting list. The detection of high heteroplasmy level pathogenic variants of undetermined clinical relevance was mostly accounted for by incomplete penetrance, such as variants conveying a risk of LHON.^9^ This adds to our understanding of how frequently secondary findings can be expected when routinely analyzing the mtDNA from ES/GS data. We also detected many low heteroplasmy level pathogenic variants of undetermined clinical relevance. Due to inconsistency of the respective proband’s phenotype with reported phenotypes, we did not pursue these variants further (for example by sequencing additional tissues).

Our study provides a glimpse into the added value of searching for novel mtDNA variants in a diverse rare disease cohort, that has not been assessed in earlier studies. By stringent filtering for high-quality variant calls at low population frequency, followed by prioritization for predicted deleterious consequence and areas of mitochondrial constraint, we identified one new diagnosis and nine promising candidates in undiagnosed families with phenotypes within the MD spectrum. The majority of the candidate variants formally remain as VUS according to the mtDNA-specifications of the ACMG/AMP guidelines for variant interpretation^28^ and additional evidence of pathogenicity is required to reach P/LP status, such as sequencing additional tissues, pursuing segregation studies in maternal relatives, further investigating blood and/or cerebrospinal fluid metabolite profiles, brain imaging, tissue histology, and respiratory chain enzymology, as well as functional studies. Candidates have been returned to local clinical or research teams in case additional study is warranted and possible, but it is beyond the scope of this study.

The field standard for assessing the population frequency of mtDNA variants is to use the homoplasmic allele frequency^28^, provided by longstanding databases such as MITOMAP, HmtDB, and MSeqDR.^6,25,43^ The majority of pathogenic mtDNA variants are, however, heteroplasmic in nature and though homoplasmic frequencies have been highly-valuable in providing evidence for or against heteroplasmic variants that are also seen at homoplasmy, they have not provided a complete picture as they have not captured the heteroplasmic frequency.^2^ In our analyses, we leveraged recently released reference population databases (gnomAD v3 and HelixMTdb) that provide both homoplasmic and heteroplasmic frequencies from over 250,000 samples collectively, mostly depleted for severe disease.^16,21^ To stay in line with field standards, we selected to integrate homoplasmic frequencies into our prioritization pipeline, given there are no recommendations in the current mtDNA-specifications of the ACMG/AMP guidelines for the use or interpretation in variant classification, and that the heteroplasmic frequency of numerous pathogenic variants exceeds the standard threshold of <1:50,000.^16^ We used the newly reported heteroplasmic frequencies and maximum observed heteroplasmy levels in reference populations to guide careful downstream clinical evaluation of candidates, with the expectation that heteroplasmic variants with a deleterious consequence on mitochondrial function are unlikely to be tolerated at high heteroplasmy levels in a reference population depleted of severe early-onset disease. We also prioritized novel variants based upon mitochondrial constraint using recently developed metrics^22^ for regional and positional constraint, with a similar rationale that variants falling in areas of mitochondrial constraint are less tolerated in humans and may play a role in disease. As mitochondrial constraint metrics are available for all mtDNA positions, our analysis could also include rRNA and non-coding variants, that are otherwise challenging to interpret due to the absence of computational prediction tools for these genomic regions. In our analysis, a number of high priority novel protein-coding candidates were supported by mitochondrial constraint data, as well as the somatic mtDNA variant likely driving the congenital sideroblastic anemia phenotype of our *POLG*-proband. At this time, all of our prioritized novel rRNA and non-coding variants in constrained regions were not of high enough clinical interest, based on the proband’s phenotype, to pursue further.

There are a number of limitations to our study. First, the primary source of DNA for sequencing in our rare disease cohort is blood, where variant heteroplasmy level is typically lower than in disease affected tissues, and can further decrease over time due to rapid replication.^44^ Therefore, though mtDNA variants can be detected in the blood in the majority of patients,^45^ in particular during childhood, it is not the optimal source of DNA for MD diagnosis. The age at DNA sample collection was not available for our cohort to further understand the impact of this limitation on our analysis. From blood we cannot conclusively rule out a MD in our undiagnosed families. This underpinned our decision to include RNA-sequencing data from probands, when available. Our RNA-sequencing data is mostly from fibroblasts or muscle tissue, offering the opportunity to capture mtDNA variants in a second tissue and potentially at a higher heteroplasmy level, as well as to increase the likelihood to detect large mtDNA deletions that are mostly isolated to muscle.^7^ In two probands (1 known diagnosis, 1 candidate) RNA-sequencing supported the presence of the variant in a second tissue. Second, in most cases, we were unable to functionally validate novel candidates by gold standard methods (e.g., single fiber analysis, cybrids) due to unavailability of patient-derived tissues. Gene editing is theoretically possible for a subset of the variant types (C>T or A>G transitions) yet is highly specialized, time consuming, and challenging to pursue.^46^ We therefore hope by sharing these candidate variants we may connect with additional affected families in the future to build evidence towards pathogenic designation.

In summary, our analysis pipeline prioritized a mtDNA variant for clinical evaluation in approximately 1 per 10 probands, adding minimal additional analytical burden to nuclear genome analysis. This gave the opportunity to capture diagnostic mtDNA variants in patients that did not necessarily have a high enough clinical suspicion of MD to prompt targeted mtDNA sequencing. In our hands, mtDNA analysis resulted in the diagnosis or prioritization of a promising candidate for 0.4% (1 in 250) of undiagnosed families with diverse rare disease phenotypes.

## Data availability

Submission to ClinVar is currently in progress for mtDNA variants that were interpreted as causal in this cohort (https://www.ncbi.nlm.nih.gov/clinvar/).

## Data Availability

Genomic and phenotypic data from the Broad CMG is available via dbGaP accession numbers phs003047 (GREGoR) and phs001272 (CMG). Access is managed by a data access committee designated by dbGaP and is based on intended use of the requester and allowed use of the data submitter as defined by consent codes.

## Acknowledgements

We thank the many families who participate in these research studies to help improve genetic diagnosis. We also thank Dr. Vamsi Mootha and Dr. Melissa Walker for providing advice on approaches to evaluate candidate mtDNA variants.

## References

1. Stenton SL, Prokisch H. Genetics of mitochondrial diseases: Identifying mutations to help diagnosis. EBioMedicine. 2020;56:102784.

2. Gorman GS, Chinnery PF, DiMauro S, et al. Mitochondrial diseases. Nat Rev Dis Primers. 2016;2:16080.

3. Frazier AE, Thorburn DR, Compton AG. Mitochondrial energy generation disorders: genes, mechanisms, and clues to pathology. J Biol Chem. 2019;294(14):5386–5395.

4. Barca E, Long Y, Cooley V, et al. Mitochondrial diseases in North America: An analysis of the NAMDC Registry. Neurol Genet. 2020;6(2):e402.

5. Gorman GS, Schaefer AM, Ng Y, et al. Prevalence of nuclear and mitochondrial DNA mutations related to adult mitochondrial disease. Ann Neurol. 2015;77(5):753–759.

6. Lott MT, Leipzig JN, Derbeneva O, et al. mtDNA Variation and Analysis Using Mitomap and Mitomaster. Curr Protoc Bioinformatics. 2013;44(123):1.23.1-26.

7. Stewart JB, Chinnery PF. The dynamics of mitochondrial DNA heteroplasmy: implications for human health and disease. Nat Rev Genet. 2015;16(9):530–542.

8. Longo N. Mitochondrial encephalopathy. Neurol Clin. 2003;21(4):817–831.

9. Yu-Wai-Man P, Votruba M, Burté F, La Morgia C, Barboni P, Carelli V. A neurodegenerative perspective on mitochondrial optic neuropathies. Acta Neuropathol. 2016;132(6):789–806.

10. Stenton SL, Shimura M, Piekutowska-Abramczuk D, et al. Diagnosing pediatric mitochondrial disease: lessons from 2,000 exomes. medRxiv. Published online June 25, 2021:2021.06.21.21259171. doi:10.1101/2021.06.21.21259171

11. Gorman GS, McFarland R, Stewart J, Feeney C, Turnbull DM. Mitochondrial donation: from test tube to clinic. Lancet. 2018;392(10154):1191–1192.

12. Wortmann SB, Koolen DA, Smeitink JA, van den Heuvel L, Rodenburg RJ. Whole exome sequencing of suspected mitochondrial patients in clinical practice. J Inherit Metab Dis. 2015;38(3):437–443.

13. Poole OV, Pizzamiglio C, Murphy D, et al. Mitochondrial DNA Analysis from Exome Sequencing Data Improves Diagnostic Yield in Neurological Diseases. Ann Neurol. 2021;89(6):1240–1247.

14. Wagner M, Berutti R, Lorenz-Depiereux B, et al. Mitochondrial DNA mutation analysis from exome sequencing-A more holistic approach in diagnostics of suspected mitochondrial disease. J Inherit Metab Dis. 2019;42(5):909–917.

15. Schon KR, Horvath R, Wei W, et al. Use of whole genome sequencing to determine genetic basis of suspected mitochondrial disorders: cohort study. BMJ. 2021;375:e066288.

16. Laricchia KM, Lake NJ, Watts NA, et al. Mitochondrial DNA variation across 56,434 individuals in gnomAD. Genome Res. 2022;32(3):569–582.

17. Calabrese C, Simone D, Diroma MA, et al. MToolBox: a highly automated pipeline for heteroplasmy annotation and prioritization analysis of human mitochondrial variants in high-throughput sequencing. Bioinformatics. 2014;30(21):3115–3117.

18. Basu S, Xie X, Uhler JP, et al. Accurate mapping of mitochondrial DNA deletions and duplications using deep sequencing. PLoS Genet. 2020;16(12):e1009242.

19. Falk MJ, Pierce EA, Consugar M, et al. Mitochondrial disease genetic diagnostics: optimized whole-exome analysis for all MitoCarta nuclear genes and the mitochondrial genome. Discov Med. 2012;14(79):389–399.

20. Riley LG, Cowley MJ, Gayevskiy V, et al. The diagnostic utility of genome sequencing in a pediatric cohort with suspected mitochondrial disease. Genet Med. 2020;22(7):1254–1261.

21. Bolze A, Mendez F, White S, et al. A catalog of homoplasmic and heteroplasmic mitochondrial DNA variants in humans. *bioRxiv*. Published online June 26, 2020:798264. doi:10.1101/798264

22. Lake NJ, Ma K, Liu W, et al. Quantifying constraint in the human mitochondrial genome. Nature. Published online October 16, 2024:1–8.

23. Landrum MJ, Lee JM, Benson M, et al. ClinVar: public archive of interpretations of clinically relevant variants. Nucleic Acids Res. 2016;44(D1):D862–D868.

24. Bianco SD, Parca L, Petrizzelli F, et al. APOGEE 2: multi-layer machine-learning model for the interpretable prediction of mitochondrial missense variants. Nat Commun. 2023;14(1):5058.

25. Preste R, Vitale O, Clima R, Gasparre G, Attimonelli M. HmtVar: a new resource for human mitochondrial variations and pathogenicity data. Nucleic Acids Res. 2019;47(D1):D1202–D1210.

26. Sonney S, Leipzig J, Lott MT, et al. Predicting the pathogenicity of novel variants in mitochondrial tRNA with MitoTIP. PLoS Comput Biol. 2017;13(12):e1005867.

27. Niroula A, Vihinen M. PON-mt-tRNA: a multifactorial probability-based method for classification of mitochondrial tRNA variations. Nucleic Acids Res. 2016;44(5):2020–2027.

28. McCormick EM, Lott MT, Dulik MC, et al. Specifications of the ACMG/AMP standards and guidelines for mitochondrial DNA variant interpretation. Hum Mutat. 2020;41(12):2028–2057.

29. Morava E, van den Heuvel L, Hol F, et al. Mitochondrial disease criteria: diagnostic applications in children. Neurology. 2006;67(10):1823–1826.

30. Slomovic S, Laufer D, Geiger D, Schuster G. Polyadenylation and degradation of human mitochondrial RNA: the prokaryotic past leaves its mark. Mol Cell Biol. 2005;25(15):6427–6435.

31. Pickett SJ, Grady JP, Ng YS, et al. Phenotypic heterogeneity in m.3243A>G mitochondrial disease: The role of nuclear factors. Ann Clin Transl Neurol. 2018;5(3):333-345.

32. Ratnaike TE, Greene D, Wei W, et al. MitoPhen database: a human phenotype ontology-based approach to identify mitochondrial DNA diseases. Nucleic Acids Res. 2021;49(17):9686–9695.

33. Huang LS, Cobessi D, Tung EY, Berry EA. Binding of the respiratory chain inhibitor antimycin to the mitochondrial bc1 complex: a new crystal structure reveals an altered intramolecular hydrogen-bonding pattern. J Mol Biol. 2005;351(3):573–597.

34. Blakely EL, Yarham JW, Alston CL, et al. Pathogenic mitochondrial tRNA point mutations: nine novel mutations affirm their importance as a cause of mitochondrial disease. Hum Mutat. 2013;34(9):1260–1268.

35. Wang J, Balciuniene J, Diaz-Miranda MA, et al. Advanced approach for comprehensive mtDNA genome testing in mitochondrial disease. Mol Genet Metab. 2022;135(1):93–101.

36. Longley MJ, Ropp PA, Lim SE, Copeland WC. Characterization of the native and recombinant catalytic subunit of human DNA polymerase gamma: identification of residues critical for exonuclease activity and dideoxynucleotide sensitivity. Biochemistry. 1998;37(29):10529–10539.

37. Wanrooij S, Goffart S, Pohjoismäki JLO, Yasukawa T, Spelbrink JN. Expression of catalytic mutants of the mtDNA helicase Twinkle and polymerase POLG causes distinct replication stalling phenotypes. Nucleic Acids Res. 2007;35(10):3238–3251.

38. Maclaine KD, Stebbings KA, Llano DA, Havird JC. The mtDNA mutation spectrum in the PolG mutator mouse reveals germline and somatic selection. BMC Genom Data. 2021;22(1):52.

39. Wong LJC, Naviaux RK, Brunetti-Pierri N, et al. Molecular and clinical genetics of mitochondrial diseases due to POLG mutations. Hum Mutat. 2008;29(9):E150–E172.

40. Ducamp S, Fleming MD. The molecular genetics of sideroblastic anemia. Blood. 2019;133(1):59–69.

41. Pontikos N, Arno G, Jurkute N, et al. Genetic Basis of Inherited Retinal Disease in a Molecularly Characterized Cohort of More Than 3000 Families from the United Kingdom. Ophthalmology. 2020;127(10):1384–1394.

42. Sallevelt SCEH, de Die-Smulders CEM, Hendrickx ATM, et al. De novo mtDNA point mutations are common and have a low recurrence risk. J Med Genet. 2017;54(2):73–83.

43. Falk MJ, Shen L, Gonzalez M, et al. Mitochondrial Disease Sequence Data Resource (MSeqDR): a global grass-roots consortium to facilitate deposition, curation, annotation, and integrated analysis of genomic data for the mitochondrial disease clinical and research communities. Mol Genet Metab. 2015;114(3):388–396.

44. Grady JP, Pickett SJ, Ng YS, et al. mtDNA heteroplasmy level and copy number indicate disease burden in m.3243A>G mitochondrial disease. EMBO Mol Med. 2018;10(6). doi:10.15252/emmm.201708262

45. Raymond FL, Horvath R, Chinnery PF. First-line genomic diagnosis of mitochondrial disorders. Nat Rev Genet. 2018;19(7):399–400.

46. Shoop WK, Bacman SR, Barrera-Paez JD, Moraes CT. Mitochondrial gene editing. Nature Reviews Methods Primers. 2023;3(1):1–18.

