## Supplemental for "Mitochondrial DNA variant detection in over 6,500 rare disease families by the systematic analysis of exome and genome sequencing data resolves undiagnosed cases"

### Supplementary Materials

1. Supplementary Methods (page 2)
2. Supplementary Tables (page 5)
3. Supplementary Figures (page 6)
4. Supplementary Results (page 8)
5. Supplementary References (page 10)

#### 1. Supplementary Methods

**mtDNA variant calling and haplogroup determination.** For genome sequencing (GS), the mitochondria mode of GATK-Mutect2 was run in Terra using the "gnomad-mitochondria" WDL file available on GitHub (https://github.com/broadinstitute/gnomad-mitochondria) for SNV and small indel calling plus haplogroup determination as previously described.[1] Default parameters were used with the exception of "vaf_filter_threshold", which was set to 0.01 to flag all variants at HL<1% as "low_allele_frac", considered as homoplasmic reference. Each GS sample-level per base coverage and VCF WDL output file was analyzed downstream in Hail version 0.2.105.[2] Each GS sample-level mtDNA aligned BAM WDL output file was used as input to the MitoSAlt pipeline for large (≥50bp) mtDNA deletion calling.[3] For exome sequencing (ES) and RNA-sequencing, the MToolBox pipeline (https://github.com/mitoNGS/MToolBox) was used for SNV and small indel calling plus haplogroup determination as previously described.[4] Default installation of MToolBox was used (install.sh). In the MToolBox config file the following mandatory parameters were set: "input_type=fastq", "ref=RCRS". All other parameters remained as default. CRAM/BAM files were converted to FASTQ files using samtools prior to running the MToolBox pipeline. Each ES/RNA-seq sample-level haplogroup annotation, per base coverage, and VCF output file was analyzed downstream in Hail version 0.2.105.[2] Each ES/RNA-seq sample-level FASTQ file containing mtDNA mapped reads was used as input to the MitoSAlt pipeline for large (≥50bp) mtDNA deletion calling.[3] For all data types, large mtDNA deletions were identified with MitoSAlt (https://sourceforge.net/projects/mitosalt/).[3] Default parameters were used, with "dna = yes" for ES/GS and "dna = no" for RNA-seq, plus "enriched = yes" for all data types in the config file. These parameters indicate the source of the input data and that the reads are enriched for mtDNA mapped reads given that all input data were previously mapped to the mtDNA and depleted of nuclear reads. Each sample's MitoSAlt tab-delimited output file of deletion calls was analyzed downstream in R version 4.1.1. As the "gnomad-mitochondria" pipeline had not previously been applied for rare disease diagnostic purposes and as the MToolBox pipeline had not previously been applied to RNA-seq data, we ran analyses of variant recall comparing "gnomad-mitochondria" to MToolBox in

24 GS samples and comparing RNA-seq to GS in 100 paired samples (see **Supplemental Results**).

**mtDNA variant annotation.** Flags were added to mark low quality variant calls that were likely false positives, as follows: i) variants at positions with coverage depth <20X were flagged as "low coverage" in the respective sample in order to only retain samples expected to have an equal or lower sequencing error rate to Sanger sequencing;[5] ii) variants in reported difficult to sequence regions (M-300:316, M-513:525, and M-16182:16194) were flagged as "artifact prone site"; iii) indel stacks, defined as indels at positions that are multi-allelic across samples in our cohort and/or gnomAD v3, were flagged as "indel stack", and iv) variants with a HL below 5% were flagged as "low heteroplasmy" due to high risk of being enriched for NUMT-derived signals. In the RNA-sequencing data, all indels variants and all SNV variants outside of the protein-coding regions were flagged for removal.

All variants were annotated with the respective gene name and gene function (protein-coding, tRNA, or rRNA) or as non-coding. Variant consequence was annotated with the Ensembl Variant Effect Predictor (VEP).[6] Variants were flagged as haplogroup defining according to Phylotree and were downloaded from the "gnomad-mitochondria" GitHub resources folder ("rCRS-centered_phylo_vars_final_update.txt"). Homoplasmic and heteroplasmic allele frequencies reported in gnomAD v3[1] and HelixMTdb (https://www.helix.com/pages/mitochondrial-variant-database)[7] reference population databases were annotated in addition to the maximum observed HL. The following computational predictions were annotated based on gene function and/or variant function: i) protein-coding and tRNA variants with HmtVar scores using the HmtVar API (https://www.hmtvar.uniba.it/);[8] ii) missense variants with APOGEE2 scores, downloaded from MitImpact (https://mitimpact.css-mendel.it/);[9,10] iii) tRNA variants with MitoTIP scores downloaded from MITOMAP (https://www.mitomap.org/foswiki/bin/view/MITOMAP/MitoTipInfo),[11,12] and PON-mt-tRNA scores downloaded from (http://structure.bmc.lu.se/PON-mt-tRNA/datasets.html/).[13] Mitochondrial constraint metrics - regional constraint and mitochondrial local constraint (MLC) scores[14] - were annotated. When data were available from the mother of the proband, the transmission of the variant was annotated as follows: i) variants detected in both the mother and proband at ≥1% HL were considered transmitted and annotated as either transmitted with “clinically significant positive heteroplasmic shift” (mother HL<60%, proband HL≥60%), “neutral heteroplasmic shift” (both mother and proband HL≥60% or HL<60%), or “clinically significant negative heteroplasmic shift” (proband HL<60%, mother HL≥0.6) in the proband; ii) variants present in the proband and not detected in the mother were annotated as “presumed *de novo* or somatic” in the proband.

#### 2. Supplemental Tables

**Table S1.** MITOMAP confirmed and ClinVar P/LP variants (separate file).

**Table S2.** Counts of high-quality variants per sample and per proband for analysis by data type.

| **Data type** | **Proband samples** (n) | **Variant count per sample** (mean) | |
|---|---|---|---|
| | | **Homoplasmic** (≥95% HL) | **Heteroplasmic** (0.05≤ HL <0.95) |
| ES (Nextera) | 1,647 | 7.7 | 1.5 |
| ES (Twist) | 2,276 | 30 | 2.7 |
| GS | 2,759 | 27 | 1.9 |
| RNA-sequencing* | 124 | 13 | 2.0 |

*RNA-sequencing analysis only includes SNVs in protein-coding regions

#### 3. Supplemental Figures

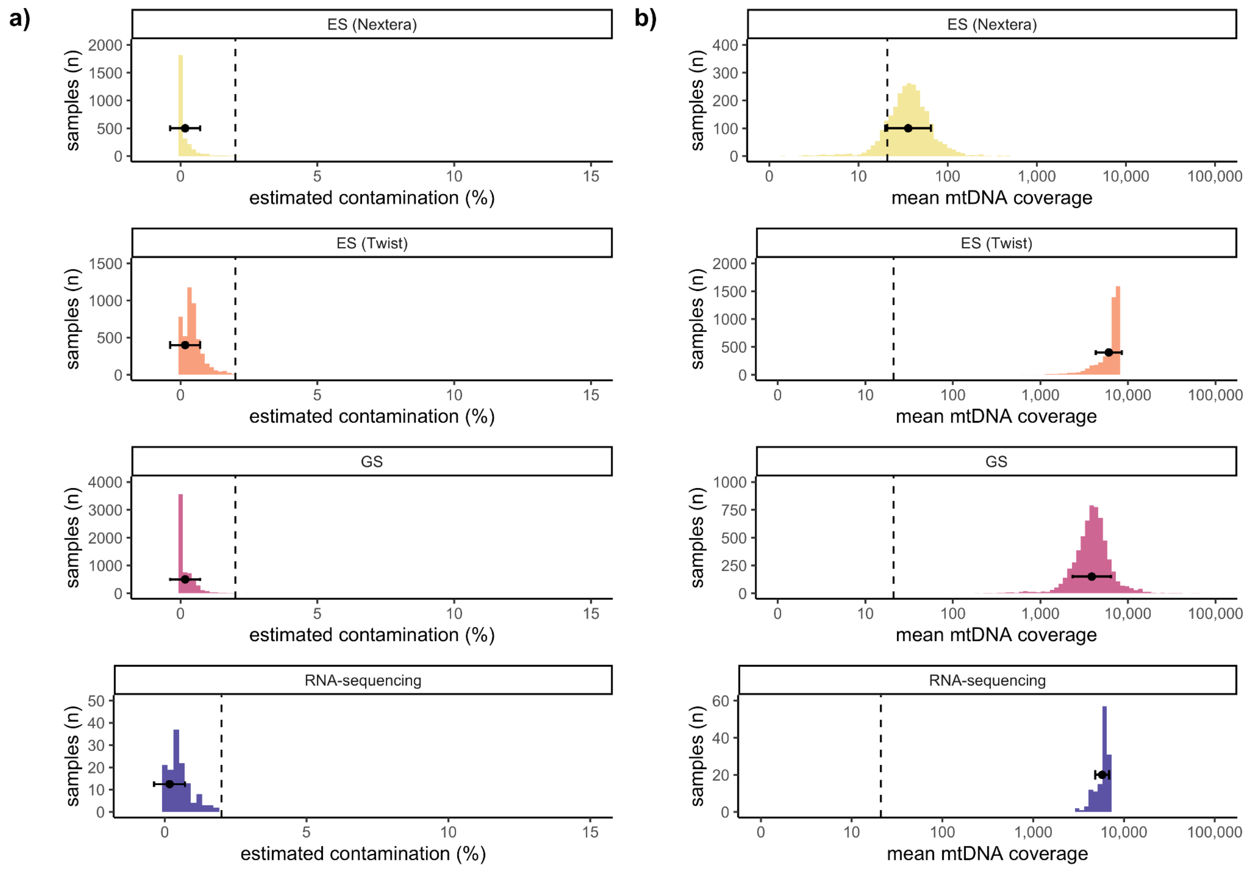


**Fig. S1. Sample level QC metrics by data type. a)** Estimated contamination (%). Samples with ≥2% contamination with high heteroplasmic haplogroup defining variants (dashed line) were excluded from the study cohort. **b)** Mean mtDNA coverage. Samples with a mean mtDNA coverage <20X (dashed line) were excluded from the study cohort. Data are displayed as histograms with corresponding mean and s.d.

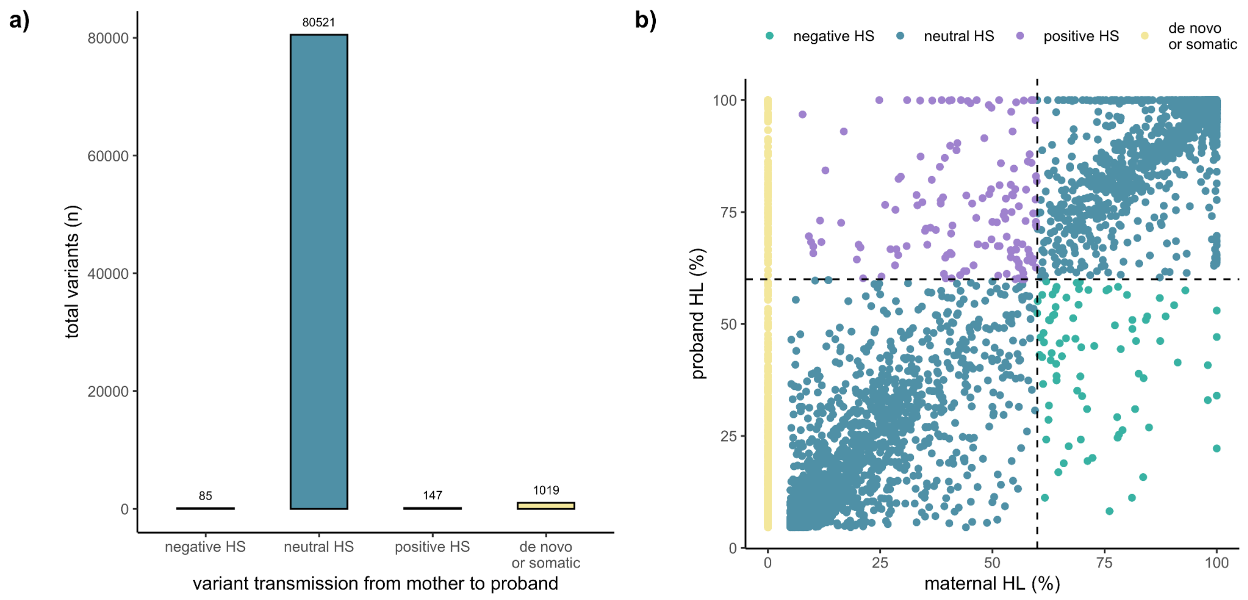


**Fig. S2. Comparison of variant heteroplasmic level in paired maternal and proband samples. a)** Total number of variants in available paired mother-proband data by variant transmission to the proband. b) Comparison of the HL of variants detected in the proband with the HL in the maternal sample, dashed lines at 60% HL indicate the typical threshold that should be exceeded to cause disease. HS, heteroplasmic shift; HL, heteroplasmy level.

#### 4. Supplemental results

**Comparison of variant calls from the "gnomad-mitochondria" pipeline and the MToolBox pipeline for 24 GS samples.** The "gnomad-mitochondria" pipeline recalled 81% (540/669) of variants detected at ≥1% HL by the MToolBox pipeline and 99% (522/526) detected at ≥5% (analysis excluded difficult to sequence regions) (**Fig. S3a**). For variants detected by both pipelines (540 variants), correlation in HL was high (R=0.99, p-value<0.0001, Pearson Correlation Coefficient) (**Fig. S3b**) and the mean difference in HL per variant was 0.06%. The pipelines agreed on the top level haplogroup for all 24 samples.

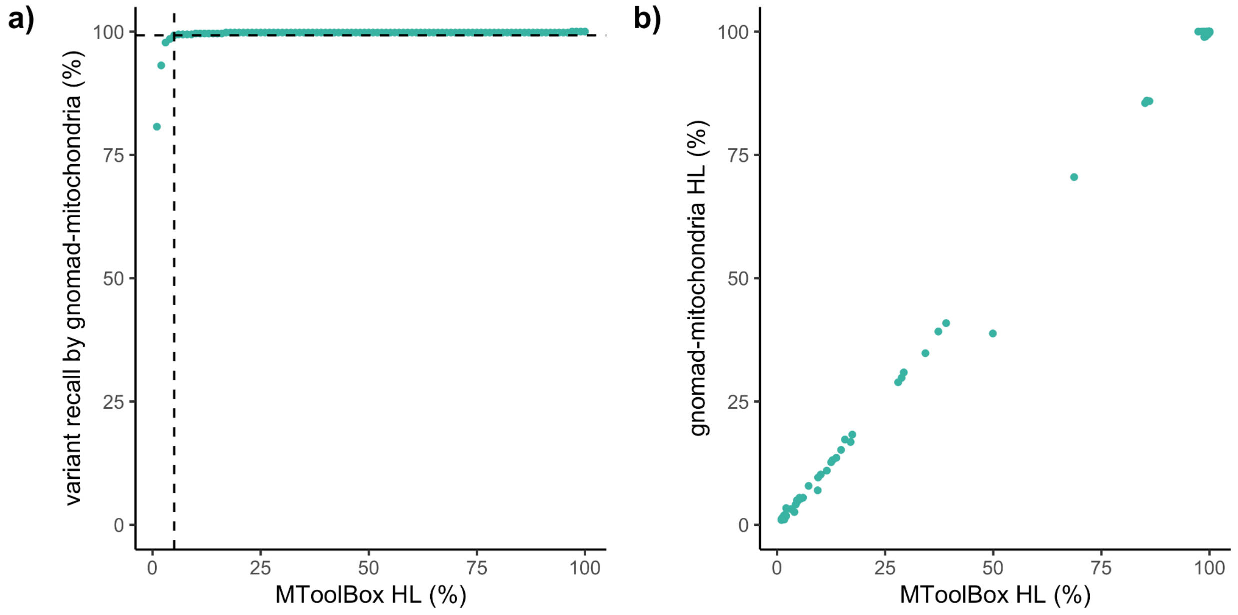


**Fig. S3. Comparison of variant calls from the "gnomad-mitochondria" pipeline and the MToolBox pipeline.**

**Comparison of variant calls from the 100 paired GS to RNA-sequencing samples.** Applying the MToolBox pipeline to RNA-sequencing data recalled 92% (1,295/1,408) of variants detected at ≥1% HL in the corresponding GS data and 97% (1,283/1,323) detected at ≥5% (analysis excluded difficult to sequence regions, indels, and non-coding regions) (**Fig. S4a**). For variants detected by both pipelines (1,295 variants), correlation in HL was high (R=0.99, p-value<0.0001, Pearson Correlation Coefficient) (**Fig. S4b**) and the mean difference in HL per variant was 1.2%. The data types agreed on the top level haplogroup for 98/100 (98%) samples. The remaining two RNA-sequencing samples had missing values for top level haplogroup.

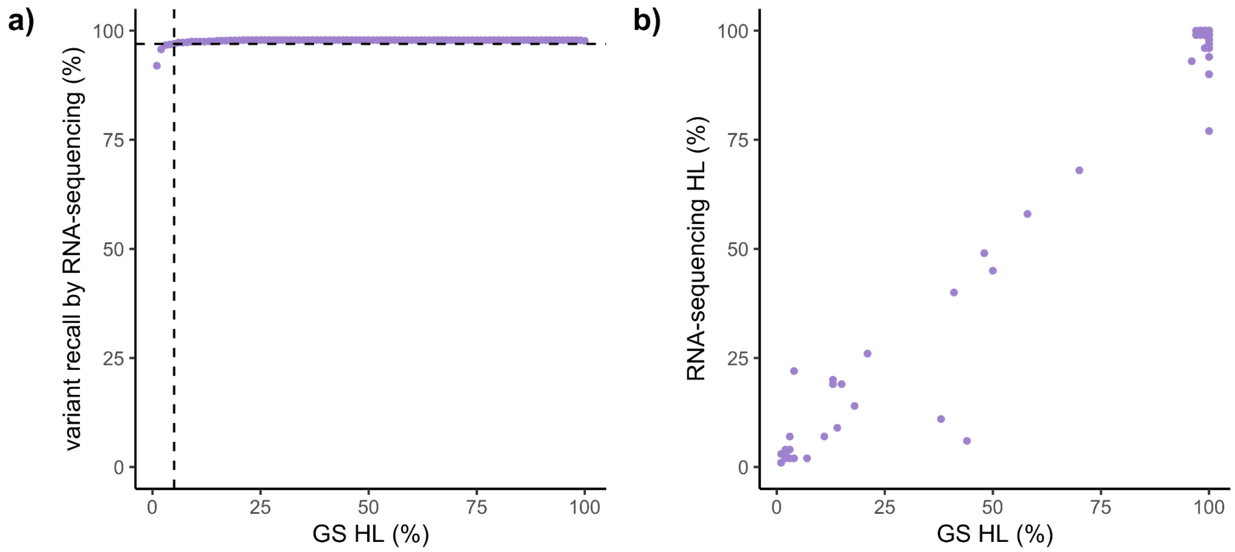


**Fig. S4. Comparison of variant calls from the 100 paired GS to RNA-sequencing samples.**

#### 5. Supplemental references


1. Laricchia KM, Lake NJ, Watts NA, et al. Mitochondrial DNA variation across 56,434 individuals in gnomAD. *Genome Res*. 2022;32(3):569-582.

2. Hail Team. Hail 0.2.105-acd89e80c345. GitHub.

3. Basu S, Xie X, Uhler JP, et al. Accurate mapping of mitochondrial DNA deletions and duplications using deep sequencing. *PLoS Genet*. 2020;16(12):e1009242.

4. Calabrese C, Simone D, Diroma MA, et al. MToolBox: a highly automated pipeline for heteroplasmy annotation and prioritization analysis of human mitochondrial variants in high-throughput sequencing. *Bioinformatics*. 2014;30(21):3115-3117.

5. Griffin HR, Pyle A, Blakely EL, et al. Accurate mitochondrial DNA sequencing using off-target reads provides a single test to identify pathogenic point mutations. *Genet Med*. 2014;16(12):962-971.

6. McLaren W, Gil L, Hunt SE, et al. The Ensembl Variant Effect Predictor. *Genome Biol*. 2016;17(1):122.

7. Bolze A, Mendez F, White S, et al. A catalog of homoplasmic and heteroplasmic mitochondrial DNA variants in humans. *bioRxiv*. Published online June 26, 2020:798264. doi:10.1101/798264

8. Preste R, Vitale O, Clima R, Gasparre G, Attimonelli M. HmtVar: a new resource for human mitochondrial variations and pathogenicity data. *Nucleic Acids Res*. 2019;47(D1):D1202-D1210.

9. Castellana S, Fusilli C, Mazzoccoli G, et al. High-confidence assessment of functional impact of human mitochondrial non-synonymous genome variations by APOGEE. *PLoS Comput Biol*. 2017;13(6):e1005628.

10. Bianco SD, Parca L, Petrizzelli F, et al. APOGEE 2: multi-layer machine-learning model for the interpretable prediction of mitochondrial missense variants. *Nat*

*Commun*. 2023;14(1):5058.

11. Sonney S, Leipzig J, Lott MT, et al. Predicting the pathogenicity of novel variants in mitochondrial tRNA with MitoTIP. *PLoS Comput Biol*. 2017;13(12):e1005867.

12. Lott MT, Leipzig JN, Derbeneva O, et al. mtDNA Variation and Analysis Using Mitomap and Mitomaster. *Curr Protoc Bioinformatics*. 2013;44(123):1.23.1-26.

13. Niroula A, Vihinen M. PON-mt-tRNA: a multifactorial probability-based method for classification of mitochondrial tRNA variations. *Nucleic Acids Res*. 2016;44(5):2020-2027.

14. Lake NJ, Ma K, Liu W, et al. Quantifying constraint in the human mitochondrial genome. *Nature*. Published online October 16, 2024:1-8.
